# A common 1.6 Mb Y-chromosomal inversion predisposes to subsequent deletions and severe spermatogenic failure in humans

**DOI:** 10.1101/2020.12.08.20245928

**Authors:** Pille Hallast, Laura Kibena, Margus Punab, Elena Arciero, Siiri Rootsi, Marina Grigorova, Rodrigo Flores, Mark A. Jobling, Olev Poolamets, Kristjan Pomm, Paul Korrovits, Kristiina Rull, Yali Xue, Chris Tyler-Smith, Maris Laan

## Abstract

Male infertility is a prevalent condition, concerning 5-10% of men. So far, only some recurrent genetic factors have been described as confident contributors to spermatogenic failure. Here, we report the first re-sequencing study of the Y-chromosomal *Azoospermia Factor c* (*AZFc*) region combined with gene dosage and Y-haplogroup determination. In analysing 2,324 Estonian men, we uncovered a novel structural variant as a high-penetrant risk factor to male infertility. The Y lineage R1a1-M458, reported at >20% frequency in several European populations, carries a fixed ∼1.6 Mb long *r2/r3* inversion destabilizing the *AZFc* region and predisposing to recurrent microdeletions. Such complex rearrangements were significantly enriched among severe oligozoospermia cases. The carrier *vs* non-carrier risk to spermatogenic failure was increased 8.6-fold (*p* = 6.0 × 10^−4^). The finding contributes to improved molecular diagnostics and clinical management of infertility. Carrier identification in young age will facilitate timely counselling and reproductive decision-making.

## Introduction

The diagnosis of male factor infertility due to abnormal semen parameters concerns ∼10% of men (*1, 2*). In today’s andrology workup ∼60% of patients with spermatogenic failure remain idiopathic (*3*). Among the known causes, the most widely considered genetic factors are karyotype abnormalities (up to 17% of patients) and recurrent *de novo* microdeletions of the Y-chromosomal *AZFa* (∼0.8 Mb), *AZFb* (∼6.2 Mb) and *AZFc* (∼3.5 Mb) regions (2–10%)(*3-5*). For more than 15 years, testing for *AZF* deletions has been strongly recommended in the diagnostic workup for infertility patients with sperm concentration <5 x 10^6^/ml (*6, 7*). Most deletion carriers represent patients with either azoospermia (no sperm) or cryptozoospermia (>0–1 million sperm/ejaculate) (*3, 8, 9*). The most prevalent deletion type is *AZFc* (∼80%), followed by the loss of *AZFa* (0.5–4%), *AZFb* (1–5%) and *AZFbc* (1–3%) regions (**Figure 1A**). Excess of recurrent *AZFc* deletions is promoted by the region’s complex genomic structure comprised of long direct and inverted amplicons of nearly identical DNA segments that lead to aberrant meiotic rearrangements in gametogenesis (*10, 11*) (**Figure 1B**). The *AZFc* full deletions remove all the multi-copy *DAZ (deleted in azoospermia 1), BPY2 (basic charge Y-linked 2*) and *CDY1* (*chromodomain Y-linked 1*) genes that are expressed in a testis-enriched manner and considered important in spermatogenesis (**Figure 1C**).

**Figure 1.**
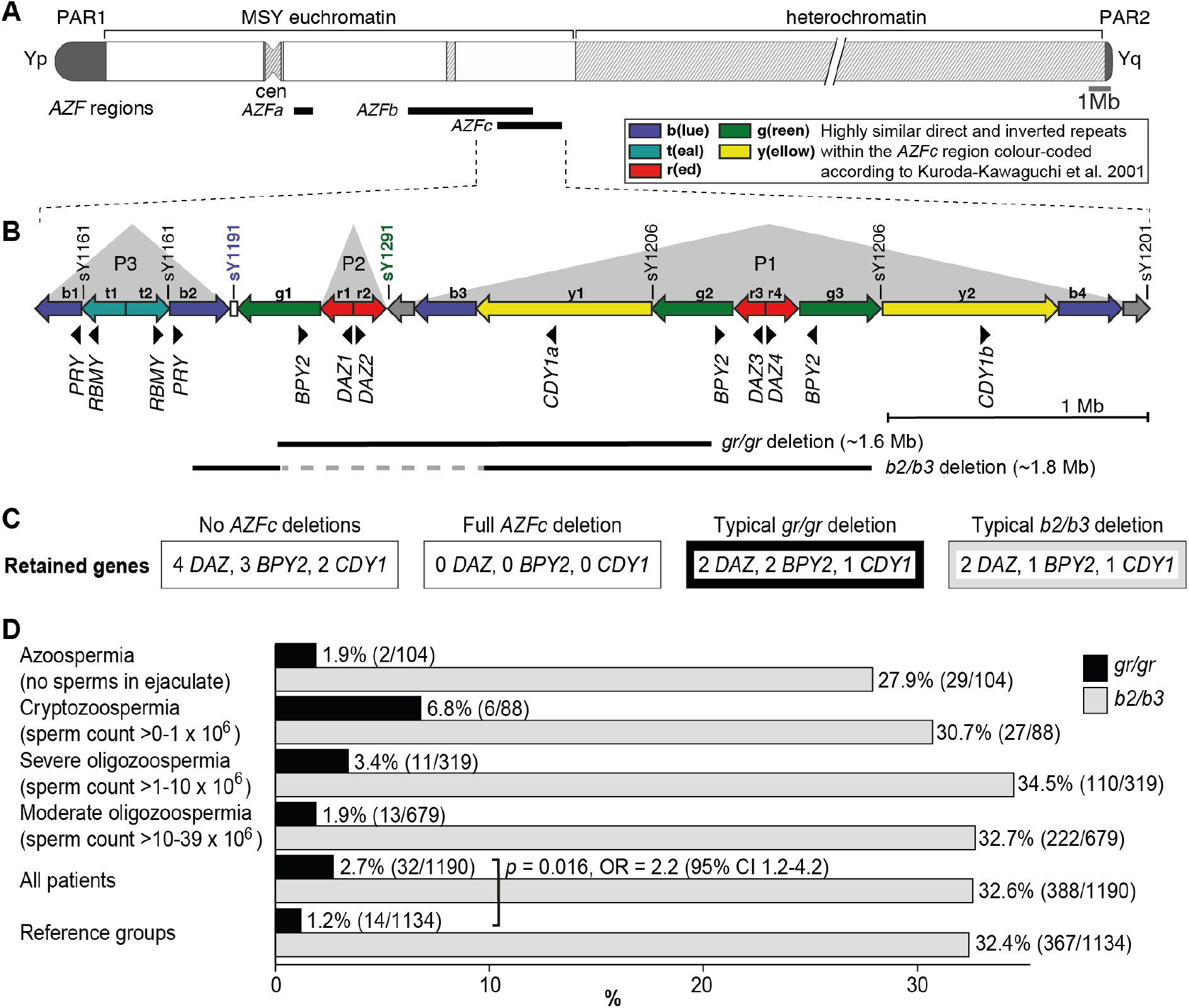
Y-chromosomal *AZFc* region and its partial deletions in the study group. **(A)** Schematic representation of the human Y chromosome with the *AZFa, AZFb* and *AZFc* regions shown as black bars. **(B)** Magnified structure of the *AZFc* region with approximate locations of multi-copy protein-coding genes, STS (sY) markers for the detection of *AZFc* partial deletions and the span of typical *gr/gr* and *b2/b3* deletions (*10*). P1-P3 (gray triangles) denote palindromic genomic segments consisting of two ‘arms’ representing highly similar inverted DNA repeats (>99.7% sequence identity) that flank a relatively short distinct ‘spacer’ sequence. Of note, the occurrence of the *b2/b3* deletion requires a preceding inversion in the *AZFc* region and therefore its presentation on the reference sequence includes also the retained segment (gray dashed line). Full details about alternative *gr/gr* and *b2/b3* deletion types are presented in **Figure S1.** **(C)** Dosage of multi-copy genes on human Y chromosomes with or without *AZFc* deletions. **(D)** Prevalence of the *gr/gr* and *b2/b3* deletions detected in the subgroups of this study. Fisher’s exact test was used to test the statistical significance in the deletion frequencies between the groups. PAR, pseudoautosomal region; MSY, male-specific region of the Y chromosome; cen, centromere; *AZF*, azoospermia factor region.

The palindromic structure of the *AZFc* region also facilitates partial deletions that are rather frequently detected in the general population (*12-15*). The most prevalent partial deletion types, named after the involved amplicons as *g(reen)-r(ed)/g(reen)-r(ed*) (lost segment ∼1.6 Mb) and *b(lue)2/b(lue)3* (∼1.8 Mb) reduce the copy-number of *DAZ, BPY2* and *CDY1* genes by roughly 50% (**Figure 1B-C, Figure S1**). The published data on the contribution of *gr/gr* and *b2/b3* deletions to spermatogenic failure are inconsistent. In European populations, the carrier status of the *gr/gr* deletion increases a risk to low sperm counts ∼1.8-fold (*14, 16, 17*). Its more variable effect on spermatogenesis has been shown in Middle Eastern and Asian populations, where the *gr/gr* deletion is completely fixed in some Y lineages, e.g. haplogroups D2 and Q1a that are common in Japan and some parts of China (*18, 19*). In contrast, the *b2/b3* deletion appears to be a risk factor for spermatogenic impairment in several East Asian and African, but not in European or South Asian populations (*20, 21*). Notably, the *b2/b3* deletion is completely fixed in Y haplogroup N3 that has a high frequency (up to 90% in some populations) in Finno-Ugric, Baltic and some Turkic-speaking people living in Northern Eurasia (*14, 15, 22*). Thus, it is unlikely that the carriership of a *gr/gr* or *b2/b3* deletion *per se* has an effect on male fertility potential. It has been proposed that this broad phenotypic variability may be explained by the diversity of *gr/gr* and *b2/b3* deletion subtypes (*23*). Y chromosomes carrying partial *AZFc* deletions may differ for the content, dosage or genetic variability of the retained genes, the overall genetic composition reflected by phylogenetic haplogroups or the presence of additional structural variants. Only limited studies have analyzed the subtypes of *gr/gr* or *b2/b3* deletions and no straightforward conclusions have been reached for their link to spermatogenic failure (*17, 24, 25*).

The current study represents the largest in-depth investigation of *AZFc* partial deletions in men recruited by a single European clinical center. We analysed 1,190 Estonian idiopathic patients with male factor infertility in comparison to 1,134 reference men from the same population, including 810 subjects with sperm parameter data available. Y chromosomes carrying *gr/gr* or *b2/b3* deletions were investigated for additional genomic rearrangements, Y-chromosomal haplogroups, dosage and sequence variation of the retained *DAZ, BPY2* and *CDY* genes. The study aimed to determine the role and contribution of *gr/gr* and *b2/b3* deletion subtypes in spermatogenic failure and to explore their potential in the clinical perspective.

## Results

### Enrichment of *gr/gr* deletions in Estonian idiopathic infertile men with reduced sperm counts

The study analyzed 1,190 Estonian men with idiopathic infertility (sperm counts 0 – 39 x10^6^/ejaculate) and a reference group comprised of 1,134 Estonian men with proven fatherhood (n=635) or representing healthy young men (n=499) (**Table 1, Table S1**). For all 2,324 study subjects, complete *AZFa, AZFb* and *AZFc* deletions were excluded.

**Table 1.**
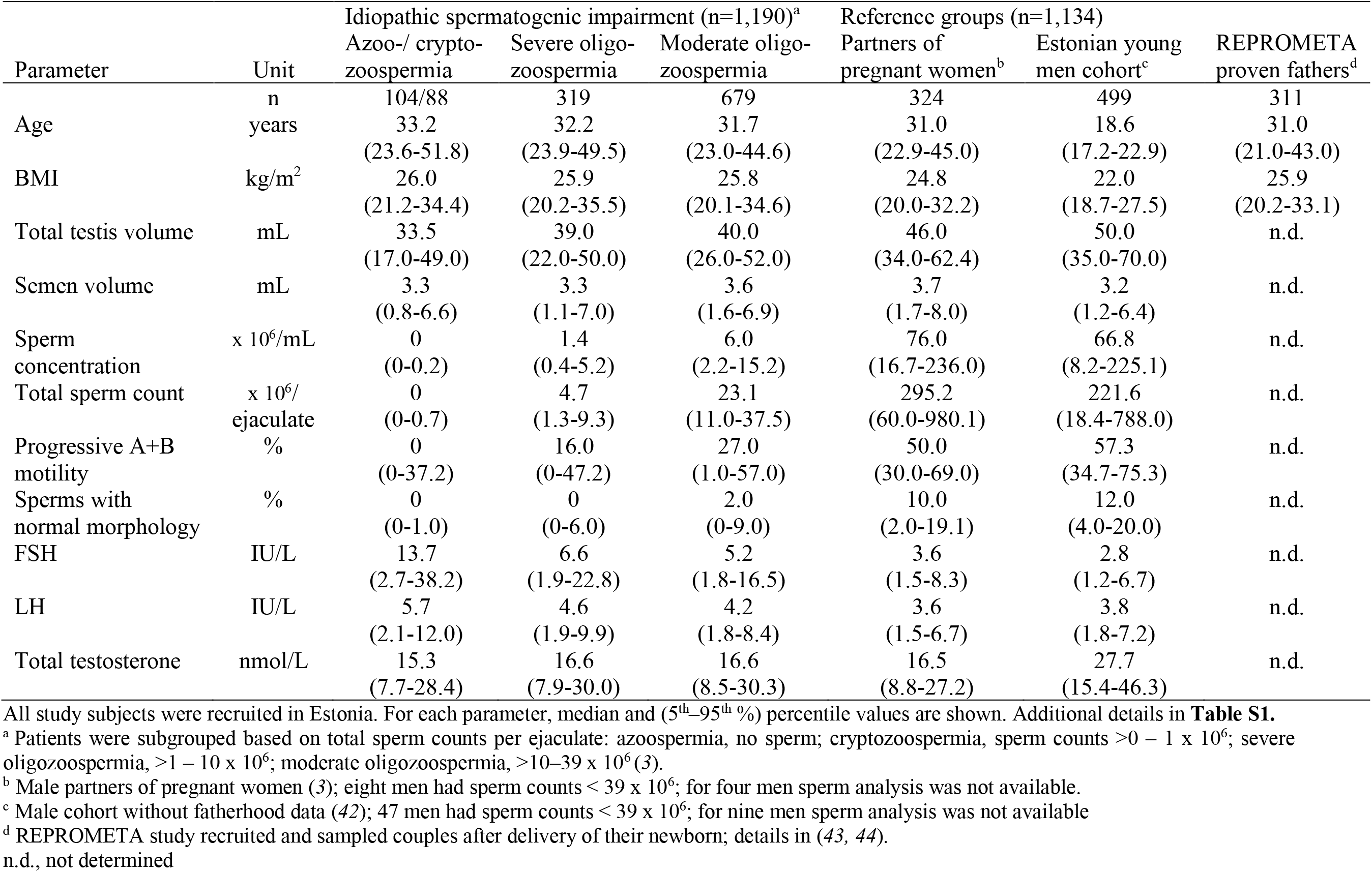
Characteristics of the patients with male factor infertility and reference groups used for comparison.

The partial *AZFc* deletions identified were *gr/gr* (n=46), *b2/b3* (n=756) and *b1/b3* (n=1, reference case) (**Table 2**). A statistically significant excess of *gr/gr* deletions was detected in idiopathic male infertility patients (2.7%; n=32/1,190) compared to reference cases (1.2%; n=14/1,134) (Fisher’s exact test, *p*=0.016; OR = 2.2 [95% CI 1.2 - 4.2]) (**Figure 1D, Table S2**). The highest frequency of *gr/gr* deletion carriers (6.8%, n=6/88) was detected in cryptozoospermia cases (sperm count >0 – 1 x 10^6^/ejaculate). However, in the reference group andrological parameters of men with or without the *gr/gr* deletion did not differ (**Table S3**). All 10 reference men with the *gr/gr* deletion and available andrological data were normozoospermic (220.3 [74.2-559.0] x 10^6^ sperm/ ejaculate). Also their other andrological parameters were within the normal range, overlapping with those of the subjects without a *gr/gr* deletion.

**Table 2.** Summary of the identified Y-chromosomal *AZF* deletion subtypes.

| Y-chromosomal rearrangements | Idiopathic male infertility patients (n) | Reference men (n) |
| --- | --- | --- |
| All analyzed cases | 1,190 | 1,134 |
| Any *AZF*c *gr/gr* deletion | 32 (2.7%) | 14 (1.2%) |
|  | Fisher's exact test, *p* = 0.016; OR = 2.2 [95% CI 1.2–4.2] |  |
| Any *AZF*c *b2/b3* deletion | 388 (32.6%) | 367 (32.4%) |
| Other type of *AZF* deletion | Loss of *b2/b3* marker<br>sY1191 (1 case) | *AZF*c *b1/b3* del (1 case);<br>partial *AZF*a del (1 case) |
| No deletion | 769 (64.6%) | 751 (66.2%) |
| *Simple partial AZFc deletions* |  |  |
| Typical *gr/gr* deletion | 19/31 (61.3%) | 8/13 (61.5%) |
| Typical *b2/b3* deletion<sup>a</sup> | 300/382 (78.5%) | 210/249 (84.3%) |
| *AZF*c partial deletion followed by *b2/b4* duplication |  |  |
| *gr/gr* del + *b2/b4* dupl<sup>b</sup> | 2/31 (6.5%) | 3/13 (23.1%) |
|  | Fisher's exact test, *p* = 0.144; OR = 0.2 [95% CI 0.0–1.6] |  |
| *b2/b3* del + *b2/b4* dupl<sup>a,b</sup> | 78/382 (20.4%) | 34/249 (13.7%) |
|  | Fisher's exact test, *p* = 0.026; OR = 1.6 [95% CI 1.0–2.4] |  |
| *AZF*c partial deletion and atypical genomic rearrangements<sup>c</sup> |  |  |
| *gr/gr* del + extra gene copies | 1 | 0 |
| *b2/b3* del + extra gene copies | 3 | 4 |
| *Complex events on the Y lineage R1a1-M458 with the preceding AZFc r2/r3 inversion* |  |  |
| *r2/r3* inv + *gr/gr* del | 8 | 2<sup>d</sup> |
| *r2/r3* inv + *gr/gr* del + *b2/b4* dupl | 1 | 0 |
| *r2/r3* inv + loss of marker sY1191 | 1 | 0 |
| + secondary gene duplications<sup>e</sup> |  |  |
| *r2/r3* inv + *b2/b3* del + *b2/b4* dupl | 0 | 1<sup>f</sup> |
| *Carriers of any AZFc gr/gr deletion type without the preceding r2/r3 inversion* |  |  |
| *gr/gr* del w/o detected *r2/r3* inv | 23/1,190 (1.9 %) | 12/1,134 (1.1%) |
|  | Fisher's exact test, *p* = 0.090; OR = 1.8 [95% CI 0.9–3.7] |  |

<sup>a</sup> Deletion subtype analysis was carried out for cases with available sufficient quantities of DNA. REPROMETA subjects were excluded from the *b2/b3* deletion subtype analysis and subsequent statistical testing due to missing andrological data.
<sup>b</sup> One or more amplicons of the retained '2x*DAZ*, 2x*BPY2*, 1x*CDY1*' (*gr/gr* deletion) or '2x*DAZ*, 1x*BPY2*, 1x*CDY1*' (*b2/b3* deletion) genes
<sup>c</sup> additional copies of *DAZ*, *BPY2* and/or *CDY1* genes inconsistent with the full '*b2/b4*' duplication
<sup>d</sup> Including one REPROMETA man without andrological data
<sup>e</sup> detected gene copy numbers 6x*DAZ*, 4x*BPY2*, 3x*CDY1*; the obligate presence of *r2/r3* inversion was defined based on Y-chromosomal phylogeny as the man carries Y lineage R1a1a1b1a1a1c-CTS11962.1 that was also identified in two cases with the *r2/r3* inversion (**Table S11**).
<sup>f</sup> Man from 'Partners of pregnant women' cohort with sperm concentration $12 \times 10^6$ /ml below normozoospermia threshold ( $15 \times 10^6$ /ml) and sperm counts $39.4 \times 10^6$ /ejaculate at the borderline of the lowest reference value ( $39.0 \times 10^6$ /ejaculate)

The patient and the reference groups exhibited similar prevalence of *b2/b3* deletions (388/1190, 32.6% *vs*. 367/1134, 32.4%; Fisher’s exact test, *p*=0.8). No apparent clinically meaningful genetic effects on andrological parameters were observed in either of the study groups (**Table S3-S4**).

### Significant overrepresentation of Y lineage R1a1-M458 in *gr/gr* deletion carriers

The Y-chromosomal haplogroups determined in 31 patients and 13 reference men carrying a *gr/gr* deletion represented 20 different lineages (patients, 17; reference men, 10; **Figure 2A, Table S5a**). Combining the phylogenetic context with the data on exact missing *DAZ* and *CDY1* gene copies (*see below*) revealed that the *gr/gr* deletion events in 44 analyzed cases must have independently occurred at least 26 times. About two-thirds of these Y chromosomes belonged to haplogroup R1, whereas the rest represented A1b, G, I and J lineages. Notably, there was a highly significant overrepresentation of Y chromosomes belonging to lineage R1a1-M458 in the *gr/gr* deletion carriers compared to the known Estonian population frequency (22.7% *vs* 5.1%; Fisher’s exact test, *p*=5.3 x 10^−4^, OR = 5.5 [95% CI 2.2-13.7]; **Figure 2B, Table S5b)** (*26*).

**Figure 2.**
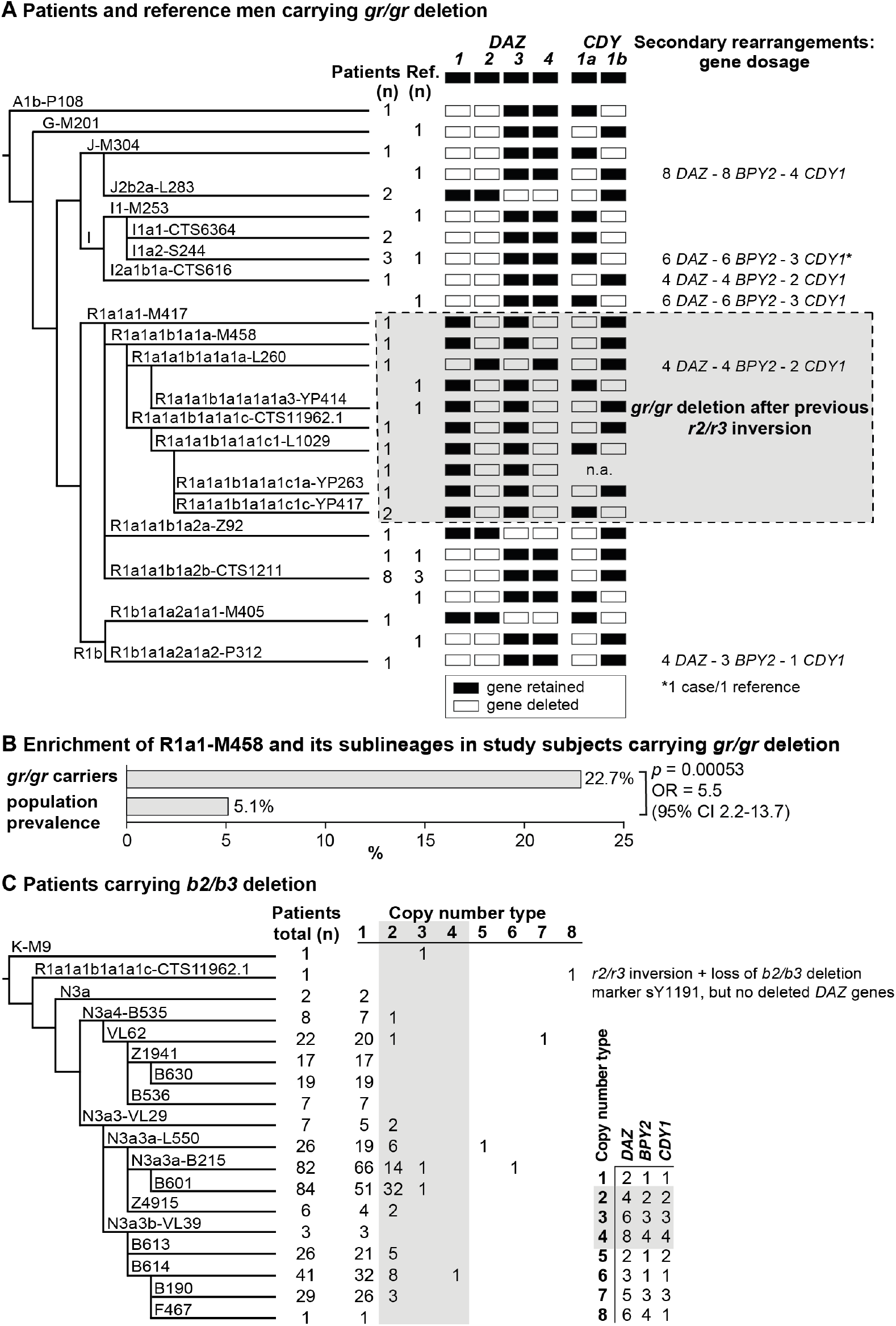
Phylogenetic relationships and gene copies in study subjects with partial *AZFc* deletions. **(A)** Y-chromosomal lineages indicated with typed terminal markers (left), deleted (white)/retained (black) *DAZ* and *CDY1* gene copies (middle) and secondary rearrangements in the *AZFc* region (right) of idiopathic male factor infertility (n=31) and reference cases (n=13) carrying the *gr/gr* deletion. The human Y-chromosomal reference sequence has four *DAZ* and two *CDY1* copies; the retained gene copies on each Y chromosome with a *gr/gr* deletion are shown as filled boxes. Chromosomes carrying atypical *gr/gr* subtypes with the loss of either the *DAZ1/DAZ3* or *DAZ2/DAZ4* gene pair due to complex genomic rearrangement combining the previous *r2/r3* inversion with a subsequent *gr/gr* deletion, are highlighted with a dashed gray square. **(B)** Enrichment of the Y-chromosomal lineage R1a1-M458 and its sub-lineages in study subjects carrying the *gr/gr* deletion in comparison to the Estonian general population (data from (*26*)). Fisher’s exact test was used to test the statistical significance between the groups. **(C)** Y-chromosomal lineages indicated with typed terminal markers (left) and the copy number of the *DAZ, BPY2* and *CDY1* gene copies (right) determined for 382 idiopathic male factor infertility cases carrying the *b2/b3* deletion. The light gray box denotes *DAZ, BPY2* and *CDY1* gene dosage consistent with full *b2/b4* duplication(s). The legend for the deletion subtype is shown in the bottom right corner. Further information on the distribution of Y-chromosomal lineages in the carriers of *AZFc* partial deletions are provided in **Tables S5-S6** and the *AZFc* rearrangement types are detailed in **Figure S1** and **Tables S7-S11**. n, number; n.a., not available; Ref, reference cases

Nearly all (99.4%) Estonian cases with the *b2/b3* deletion belonged to the Y haplogroup N3, in which this event is fixed (*13, 15*). The most commonly detected sub-lineage was N3a3a-L550 (∼51% of 436 typed chromosomes) and in total 15 different haplogroups that had diverged after the *b2/b3* deletion event in the common ancestor of N3 were present in Estonian men (**Figure 2C, Table S6**). *b2/b3* Y chromosomes representing non-N3 lineages were detected in two patients and three reference men. Lineage typing was possible for three of them, who carried either K-M9 (one patient) or R1a1a1b1a1a1c-CTS11962.1 (one patient and one reference case).

### Increased prevalence of *b2/b3* deletion followed by *b2/b4* duplication in infertile men

The expected retained copy number of *DAZ, BPY2* and *CDY1* genes consistent with the typical *gr/gr* deletion was found in 37/44 (∼84%) cases (**Figure 2A, Table 2, Table S7)**. Three patients and three reference men carried a secondary *b2/b4* duplication adding one or more amplicons of [two *DAZ* – two *BPY2* – one *CDY1*] genes with no apparent effect on infertility status (Fisher’s exact test, *p*=0.34). Notably, four of six samples with secondary *b2/b4* duplication events were identified in haplogroup I. Similarly, 78.5% of patients and 84.3% reference men with the *b2/b3* deletion presented gene dosage consistent with the typical deletion (**Figure 2C, Table 2, Table S7-S8**). Indicative of recurrent secondary events, one or more *b2/b4* duplications of [two *DAZ* – one *BPY2* – one *CDY1*] genes were identified in 13 haplogroups, including non-N3 lineages K-M9 and R1a1a1b1a1a1c-CTS11962.1. Although secondary *b2/b4* duplications were detected with significantly higher prevalence in patients compared to the reference men (n=78/382, 20.4% vs n=35/249, 14.1%; Fisher’s exact test, *p*=0.026, OR = 1.57 [95% CI 1.02 - 2.42]), no consistent effect of increased gene copy number on andrological parameters was observed (**Tables S3-S4**). Reference men with *b2/b4* duplication compared to subjects with no *AZFc* rearrangements showed a trend for lower FSH (median 2.3 [5-95% range 1.4-7.5] *vs* 3.2 [1.3-7.1] IU/L; *P*<0.05) and LH (3.1 [1.7-5.0] *vs* 3.8 (1.7-7.2) IU/L; *P*<0.05). Additionally, in eight subjects with *AZFc* partial deletions, further atypical Y-chromosomal genomic rearrangements were detected, but also with no clear evidence for a phenotypic effect (**Table 2, Table S7**).

The data gathered from this analysis thus suggest that the dosage of *DAZ, BPY2* and *CDY1* genes does not play a major role in modulating the pathogenic effect of the *gr/gr* and *b2/b3* deletions.

### No specific *DAZ* or *CDY1* gene copy is lost in men with spermatogenic failure

The major *b2/b3* deletion subtype in both patients (99.7%) and reference cases (98.1%) was the loss of *DAZ3*-*DAZ4-CDY1a* genes, whereas the most frequent *gr/gr* deletion subtypes were the loss of *DAZ1-DAZ2-CDY1a* (41.9%, 18/43 cases) and *DAZ1-DAZ2-CDY1b* (25.6%, 11/43 cases) combinations (**Figure 2A, Table S9-S10**). The observed prevalence of the major *gr/gr* subtypes was concordant with the published data on other European populations (42.5% and 25.5%, respectively; (*17*)). As these *gr/gr* deletion subtypes are prevalent in the reference group (total 11 of 13, 84.6%), their major role in spermatogenic impairment can be ruled out. As a novel insight, a subset of these Y chromosomes showed lineage-specific loss of some exon 7 subtypes of the retained *DAZ4* gene (**Figure S2; Table S10**). All five exons 7Y were missing in the Y chromosomes with the *DAZ1-DAZ2-CDY1a* deletion that had occurred in sub-lineages of the R1a1a1b1a2 haplogroup (9/31 patients, 3/13 reference men). The exon 7F was lost in haplogroup I1 and its sub-lineages (5/31, 2/13) carrying the *DAZ1-DAZ2-CDY1b* deletion. There was no evidence that loss of *DAZ4* exons 7Y or 7F has any phenotypic consequences. Most likely, this observation reflects gene conversion events from *DAZ3* to *DAZ4* as the former lacks both, exons 7Y and 7F.

Taken together, our findings indicate that neither the loss of the *DAZ1-DAZ2* nor the *DAZ3-DAZ4* gene pair, combined with either a *CDY1a* or *CDY1b* gene, directly causes spermatogenic failure. Interestingly, no Y chromosomes were observed with fewer than two retained *DAZ* genes.

### Y lineage R1a1-M458 carries a fixed *r2/r3* inversion predisposing to recurrent deletions

Novel atypical *gr/gr* and *b2/b3* deletion subtypes with the loss of an unusual *DAZ* gene pair were identified (**Figure 2A, Table 2, Table S9-S11**). Eight patients and two reference cases with a *gr/gr* deletion were missing *DAZ2-DAZ4* genes. Loss of *DAZ1-DAZ3* genes followed by a subsequent *b2/b4* duplication event was identified in one infertile and one reference case with either *gr/gr* or *b2/b3* deletion, respectively. All but one subject with this atypical pair of lost *DAZ* genes belonged to the Y haplogroup R1a1-M458 and its sub-lineages, significantly enriched in *gr/gr* deletion carriers (**Figure 2B, Table S5b**). The most parsimonious explanation to explain the simultaneous deletion of either *DAZ1-DAZ3* or *DAZ2-DAZ4* genes is a preceding ∼1.6 Mb long inversion between the *r(ed)2* and *r(ed)3* amplicons (**Figure 3A**). This new inverted structure might be more susceptible to recurrent deletions as it has altered the internal palindromic structure of *AZFc* region. In *r2/r3* inversion chromosomes, the largest palindrome P1 is almost completely lost and the size of the palindrome P2 is greatly expanded by positioning the homologous *g1/g2* segments in an inverted orientation. The *r2/r3* inversion is consequently expected to destabilize the *AZFc* region as several long DNA amplicons with highly homologous DNA sequence are positioned in the same sequence orientation (*b2, b3* and *b4; g2* and *g3; y1* and *y3*). Therefore, they are prone to non-allelic homologous recombination mediating recurrent deletions and duplications. Since these atypical deletion subtypes were identified only in a specific Y-chromosomal haplogroup, the detected *r2/r3* inversion must have occurred only once in the common ancestor of R1a1-M458 sub-lineages. One patient with the loss of *DAZ2-DAZ4* carried haplogroup R1a1a1-M417, an ancestral lineage to R1a1-M458 (**Figure 2A**). However, lineage R1a1a1-M417 is not fixed for this inversion since its other sub-lineage, R1a1a1b1a2, does not carry it and any subsequent inversion restoring the exact original *AZFc* structure is not credible. The more parsimonious explanation is that the inversion occurred in a sub-lineage of R1a1a1-M417 that has to be yet determined.

**Figure 3.**
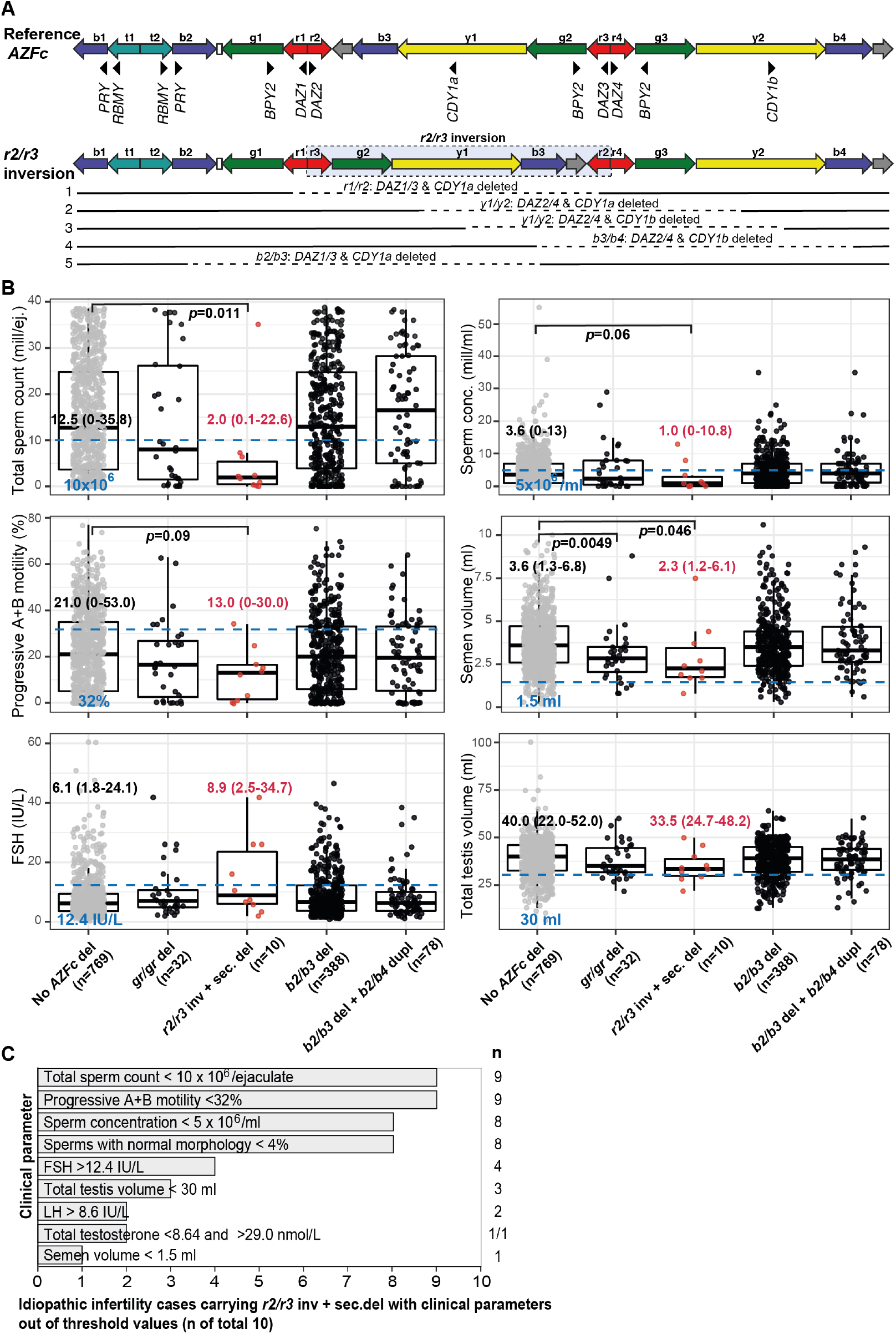
Complex structural variants at the Y-chromosomal lineage R1a1-M458 and their effect on andrological parameters. **(A)** Schematic presentation of the Y chromosome with the *r2/r3* inversion compared to the reference sequence. The *r2/r3* inversion structure nearly destroys the large palindrome P1 and, consequently, destabilizes the *AZFc* region since several long DNA amplicons with highly similar DNA sequence (*b2, b3* and *b4*; *g2* and *g3*; *y1* and *y3*) are positioned in the same sequence orientation. This structure promotes non-allelic homologous recombination mediating recurrent deletion and duplication events. The approximate regions removed by the identified *gr/gr* and *b2/b3* deletions arising on the *r2/r3* inverted Y chromosome are shown as dashed lines. **(B)** Distribution of andrological parameters in the idiopathic male factor infertility cases (total sperm counts 0 – 39 x 10^6^) subgrouped based on the structure of the *AZFc* region. The median (5–95% range) of each parameter is shown for cases carrying the *r2/r3* inversion plus secondary deletions compared to infertile men without any *AZFc* deletion. The Pairwise Wilcoxon Rank sum test was applied to estimate the statistical difference between groups. Threshold values (shown in blue) for sperm parameters corresponding to severe spermatogenic failure are based on international guidelines (*27*). For reproductive hormones, reference values of the laboratory service provider are shown. The empirical threshold for the total testis volume was based on routinely applied clinical criteria at the AC-TUH. For full details see **Table S3 and Table S11.** **(C)** The majority of idiopathic infertility cases carrying the *r2/r3* inversion plus secondary *AZFc* partial deletions (total n=10) exhibit severe oligoasthenoteratozoospemia (OAT) defined as extremely reduced sperm counts (<5 x 10^6^/ml) and concentration (<10 x 10^6^/ejaculate) combined with low fraction of sperms with normal morphology (<4% normal forms) and motility (<32% progressive motile spermatozoa). Reference values for andrological parameters have been applied as referred in sub-figure B. As total testis volume is mostly within the expected range, their infertility is not caused by intrinsic congenital testicular damage but rather due to severe spermatogenic failure *per se*. Del, deletion, inv, inversion, dupl, duplication, n, number; sec, secondary, mill, million, ej., ejaculate

Based on the Y-chromosomal phylogenetic data, one additional patient was identified as an obligate carrier of the *r2/r3* inversion as his Y chromosome represents the lineage R1a1a1b1a1a1c-CTS11962.1 that was also identified in two cases with the *r2/r3* inversion. This patient exhibited signs of unusual deletion and duplication events in the *AZFc* region as he carried six *DAZ*, four *BPY2* and three copies of the *CDY1* gene (**Figure 2B, Table 2, Table S8**).

Among the analyzed 2,324 men, 13 cases with the complex *AZFc* rearrangement combining *r2/r3* inversion with a subsequent deletion, represented 0.6% (**Table 3**). Considering the reported population prevalence of R1a1-M458 lineage in Estonians (5.1%; (*26*)), the estimated number of subjects representing this Y lineage in the study group was ∼119. Thus, approximately one in ten chromosomes with the *r2/r3* inversion had undergone a subsequent deletion event (13/119, 11%).

**Table 3.** Enrichment of the *AZFc r2/r3* inversion followed by a partial *AZFc* deletion in men with severe spermatogenic failure

| Group | All (n) | AZFc r2/r3 inversion + AZFc partial deletion |  |  |
| --- | --- | --- | --- | --- |
|  |  | Estimated non-carriers (n) | Detected carriers (n) | % of carriers in the (sub)group |
| *a. Full study group* |  |  |  |  |
| All analyzed study subjects | 2,324 | 2,311 | 13 | 0.6% |
| Study subjects with sperm counts | 2,000 | 1,988 | 12 | 0.6% |
| *Subjects stratified based on total sperm counts per ejaculate* |  |  |  |  |
| Sperm counts 0 – 10 x 10<sup>6</sup> | 524 | 515 | 9 | 1.7% |
| Sperm counts >10 x 10<sup>6</sup> | 1,476 | 1,473 | 3 | 0.2% |
| Fisher's exact test, $p = 6.0 \times 10^{-4}$, OR = 8.6 [95% CI 2.3–31.8] | | | | |
| *b. Carriers of the Y lineage R1a1a-M458<sup>a</sup>* |  |  |  |  |
| In all analyzed study subjects | 119 | 106 | 13 | 11.0% |
| In study subjects with sperm counts | 102 | 90 | 12 | 11.8% |
| *Subjects stratified based on total sperm counts per ejaculate* |  |  |  |  |
| Sperm counts 0 – 10 x 10<sup>6</sup> | 27 | 18 | 9 | 33.7% |
| Sperm counts >10 x 10<sup>6</sup> | 75 | 72 | 3 | 4.0% |
| Fisher's exact test, $p = 3.0 \times 10^{-4}$, OR = 12.0 [95% CI 2.9–48.9] | | | | |

<sup>a</sup> expected number of Y lineage R1a1-M458 in each subgroup was estimated using the known Estonian population prevalence 5.1% (26)

### *r2/r3* inversion promotes recurrent deletions that lead to severe oligoasthenoteratozoospermia

Idiopathic infertility cases carrying the *r2/r3* inversion with a subsequent deletion in the *AZFc* region (n=10) exhibited extremely low sperm counts compared to subjects without any *AZFc* deletions (median 2.0 *vs* 12.5 x 10^6^/ejaculate; Wilcoxon test, *p*=0.011) (**Figure 3B, Figure S3, Table S3)**. Nine of 10 men showed severe spermatogenic failure (total sperm counts <10 x 10^6^/ejaculate), either azoo- (n=1), crypto- (n=3) or severe oligozoospermia (n=5) (**Table S10-S11**). They also showed consistently the poorest sperm concentration (median 1.0 x 10^6^/ml) and progressive motility (13%), as well as the lowest semen volume (2.3 ml) compared to the rest of analyzed infertile men. The data suggests that extreme oligoasthenoteratozoospermia (OAT) observed in these subjects was due to the severely affected process of spermatogenesis, whereas their testicular volume and hormonal profile were within the typical range of male factor infertility cases (**Figure 3C**).

When all the men with andrological data (n=2,000) were stratified based on sperm counts, there was a highly significant enrichment of the *r2/r3* inversion with a subsequent deletion in men with severe spermatogenic failure (sperm counts 0 – 10 x 10^6^) compared to the rest (1.7% *vs* 0.2%, Fisher’s exact test, *p*=6.0 x 10^−4^, OR=8.6 [95% CI 2.3 – 31.8]; **Table 3**). The estimated number of phenotyped subjects representing the Y haplogroup R1a1-M458 with the fixed *r2/r3* inversion was 102 (based on population prevalence 5.1%; (*26*)). Among carriers of this Y lineage, 33.7% of men with sperm counts 0–10 x 10^6^ (9/27), but only 4.0% with sperm counts >10 x 10^6^ (3/72) had undergone a subsequent *AZFc* partial deletion (Fisher’s exact test, *p*= 3.0 x 10^−4^, OR = 12.0 [95% CI 2.9 - 48.9]).

Only three reference cases carried a Y chromosome with the *r2/r3* inversion with a subsequent *AZFc* partial deletion. At the time of phenotyping, all three subjects were younger (aged 18, 21 and 23 years) than the variant carriers in the idiopathic infertility group (median 32.4, range 26-51 years) (**Table S11**). The only reference subject with this complex *AZFc* rearrangement, but unaffected sperm analysis was the youngest (18 years). Notably, another reference man (23 years) with andrological data would actually be classified, based on WHO guidelines (*27*), as an oligozoospermia case (sperm concentration 12 x 10^6^/ml *vs* threshold 15 x 10^6^/ml). Also, his total sperm counts (39.4 x 10^6^/ejaculate) represented a borderline value.

### Sequence diversity of the retained *DAZ, BPY2* and *CDY* genes is extremely low and has no detectable effect on sperm parameters

The re-sequenced retained *DAZ1-4, BPY2* and *CDY1-2* genes were characterized by extremely low nucleotide variability in all Y-chromosomal lineages and deletion subtypes (**Table S12)**. For 476 samples (*gr/g*r, n=40; *b2/b3*, n= 436) re-sequenced for the >94 kb region, a total of 42 variants were identified with median 0.8 variants/kb and maximum two variants per individual. Most of them were previously undescribed (41), singletons (31) and/or non-coding SNVs/short indels (36) (**Table S13)**. The *CDY2a-CDY2b* genes harbored only one variable site, whereas *DAZ1-DAZ2* carried 24 or 26 SNVs/indels. Most variants appeared paralogous as both the reference and alternative alleles were identified. Among the four detected missense variants, CDY1b p.T419N was fixed in all three *CDY1b* copies present on the Y chromosome with the *b2/b3* deletion plus *b2/b4* duplications that represented an oligozoospermia case. However, the effect of this conservative substitution is unclear.

There was thus no evidence that the sequence variation in *DAZ, BPY2* and *CDY* genes has any effect on infertility related parameters in the subjects examined.

## Discussion

We conducted a comprehensive investigation of partial deletion subtypes of the Y-chromosomal *AZFc* region in 2,324 Estonian men, approximately half with idiopathic spermatogenic impairment (n=1,190) in comparison to the reference group (n=1,134). Importantly, 2,000 men had undergone full and uniformly conducted andrological workup at a single clinical center, facilitating fine-scale genotype-phenotype analysis. Previously, no study had undertaken re-sequencing of the retained *DAZ, BPY2* and *CDY* genes along with the assessment of the Y haplogroup, dosage and retained/deleted genes in the *gr/gr* or *b2/b3*-deleted chromosomes in both infertile men and controls. Concordant with reports from other European populations, the *gr/gr*, but not the *b2/b3* deletion, is a risk factor for spermatogenic impairment in Estonian men with >2-fold increased susceptibility to infertility (**Figure 1D**). However, the gathered data on the large group of reference men in the current study demonstrated the existence of Y chromosomes carrying a *gr/gr* deletion without any documented effect on andrological parameters (**Table S3**). As a novel finding, the study uncovered complex *AZFc* rearrangements within a specific Y haplogroup, R1a1-M458 and its sub-lineages, causing severe spermatogenic failure in the majority of carriers (**Figure 3, Table 3**). This Y lineage has undergone a ∼1.6 Mb *r2/r3* inversion in the *AZFc* region that has disrupted the structure of the palindromes P1 and P2, promoting subsequent recurrent deletions and consequently, severely impaired the process of spermatogenesis.

Consistent with key early observations (*14, 17, 23*), this study supports the recurrent nature and high subtype diversity of the *AZFc* partial losses that are currently considered jointly under the umbrella term ‘*gr/gr* deletions’. The 44 detected *gr/gr* deletions in our study sample were estimated to have originated independently at least 26 times across the Y phylogenetic tree and include seven different combinations of *DAZ* and *CDY1* gene losses. Apparently, there is a substantial undescribed heterogeneity in the spread and structure of *gr/gr* deletions that in turn results in phenotypic variability of the genetic effects. Unexpectedly, one in four Estonian *gr/gr* deletion carriers belonged to the Y-chromosomal haplogroup R1a1-M458 (and its sub-lineages) (**Figure 2A-B**; 22.7% vs 5.1% reported as the Estonian population frequency (*26*)). Notably, a previous study has reported a significant enrichment of the haplogroup R1a (ancestral lineage to the R1a1-M458) among *gr/gr*-deleted chromosomes in the Polish population (*14*), which has a high prevalence, 25%, of R1a1-M458 (*26*) (**Table S14**). All the Estonian *gr/gr* cases and also additional *b2/b3* deletion chromosomes representing this Y lineage carried unusual retained *DAZ* gene pairs (*DAZ1-DAZ3* or *DAZ2-DAZ4*) in combination with either *CDY1a* or *CDY1b* gene copy (**Figure 2**). These complex *AZFc* rearrangements were best explained by a preceding (and apparently fixed in R1a1-M458) ∼1.6 Mb inversion between the homologous *r2* and *r3* amplicons, followed by recurrent secondary partial *AZFc* deletions (**Figure 3A**). The latter are facilitated by large ampliconic segments positioned in the same orientation. Inversions in the *AZFc* region are not uncommon, but none of the previously described inversions is expected to substantially disrupt the core palindromic structure of the *AZFc* region (**Figure S1**) (*13, 23*). In contrast, the *r2/r3* inversion disrupts the structure of palindrome P1 and expands the size of the P2 palindrome more than two-fold (**Figure 3A**). The critical role of intact P1-P2 palindromes in the *AZFc* structure is supported by the observation that no Y chromosomes have been described with a single *DAZ* gene copy, whereas the inverted *DAZ* gene pairs form the ‘heart’ of both P1 and P2.

As a likely scenario, this complex *AZFc* rearrangement may predispose to spermatogenic impairment through substantial destabilization of the intra-chromosomal structure affecting meiotic recombination and chromosomal segregation. Among 12 Estonian subjects carrying the *AZFc r2/r3* inversion followed by partial deletions and with available data for sperm counts, nine cases exhibited severe spermatogenic failure, two had moderate oligozoospermia and only one case (aged 18 years) was normozoospermic (**Figure 3B-C**). This represented ∼8-9-fold enrichment of this complex rearrangement among men with severely reduced sperm counts (0 – 10 x 10^6^; **Table 3**). This genetic effect was observed specifically on the effectiveness of spermatogenesis, whereas the measurements of bitesticular volume and reproductive hormone levels did not stand out among the rest of analyzed infertile men. To our knowledge, no other Y-lineage specific risk variants for spermatogenic impairment have been reported so far. Previously, the *DAZ2-DAZ4* deletion had been shown as a high-risk factor for male infertility in the Tunisian population, but the Y haplogroups of those subjects was not investigated (*24*). The survival of such a high-risk lineage in the population seems at first sight surprising, but may be accounted for by its possible age-specific effects on spermatogenesis, which may be exacerbated by the recent general decline in sperm count (*28*). In the past, this lineage may not have been disadvantageous.

This study outcome has notable clinical implications for the improvement of molecular diagnostics and reducing the proportion of idiopathic male factor infertility cases. In Northern and Central Europe, the prevalence of R1a1-M458 haplogroup carrying the *r2/r3* inversion ranges from ∼1% in the Netherlands and Denmark to ∼2-5% in Austria, Hungary, Germany, Baltics and most Balkan countries, whereas it is widespread in Slavic populations and carried by 12-26% of men (**Figure 4, Table S14**)(*26*). In these populations, recurrent secondary *AZFc* partial deletions on Y chromosomes representing the R1a1-M458 haplogroup (and its sub-lineages) may potentially explain from 0.3% up to ∼9% of cases presenting severe spermatogenic impairment (sperm counts <10 million per ejaculate). Further studies in other populations and large samples of patients and normozoospermic controls are required to fully establish the value of extending the current recommended testing of Y-chromosomal deletions by including the analysis of this novel Y-lineage-specific pathogenic *AZFc* rearrangement (a detailed protocol is provided in **Supplemental Subjects and Methods**).

**Figure 4.**
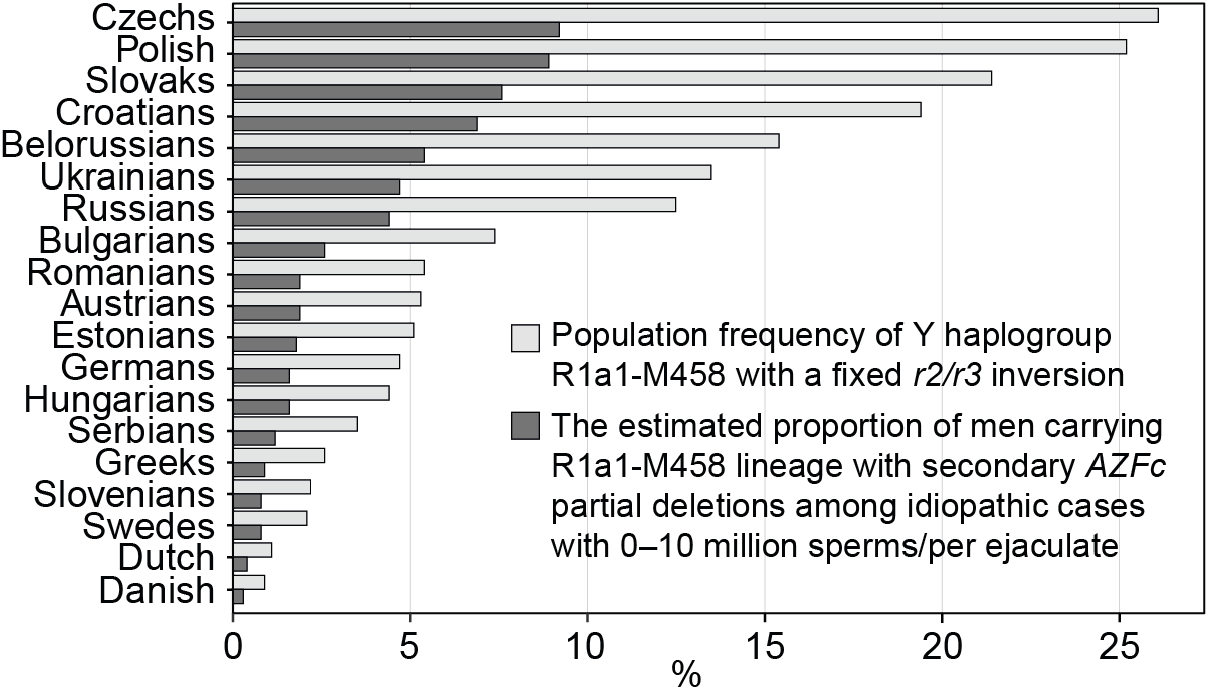
The prevalence of the Y-chromosomal haplogroup R1a1-M458 carrying a fixed *r2/r3* inversion. Population frequencies of the haplogroup were derived from (*26*). The proportion of subjects carrying this Y lineage (and its sub-lineages) and who had undergone a subsequent secondary *AZFc* deletion among idiopathic cases with severe spermatogenic failure (sperm counts 0–10 x 10^6^/ejaculate) was estimated for each population based on data from the current study in the Estonian population. For full details see **Table S14**.

The evidence from the literature has shown that the increased prevalence of either *gr/gr* or *b2/b3* deletions in infertility cases appears to be population-dependent (*16, 20*). It can be speculated that also in other populations some specific Y lineages may carry *AZFc* structural variants that in combination with partial deletions (or other rearrangements) predispose to chromosomal instability in the complex process of spermatogenesis involving multiple well-coordinated cell divisions. So far, the largest conducted study on the Y-chromosomal phylogeny of *gr/gr* deletion carriers included 152 infertile subjects representing seven countries with different population genetic structures (*17*). However, the number of cases per population was low and the study included only 17 fertile men. Also, the study did not include fine-scale analysis of Y sub-lineages and the retained gene content. Long-range re-sequencing of the whole *AZFc* region in large numbers of men would be the preferred approach to uncover its structural complexity. Additional pathogenic *AZFc* rearrangements may also exist among Estonian infertile men. When omitting the cases carrying a *gr/gr* deletion at the *r2/r3* inversion background, a non-significant enrichment of the remaining *gr/gr* deletion chromosomes can be observed in patients compared to reference men (1.9 *vs* 1.1%, *p* < 0.1; **Table 2**).

In addition to the main finding, our deep re-sequencing dataset revealed that neither the dosage, sequence variation nor exact copy of the retained *DAZ, BPY2* and *CDY1* gene showed any detectable effect on spermatogenic parameters. All chromosomes with *AZFc* partial deletions exhibit extremely low overall sequence variation of the retained *DAZ, BPY2* and *CDY* genes. This observation is consistent with previous reports showing low levels of genetic diversity of the human Y chromosome (*29*) and suggesting that novel variants may be rapidly removed by active gene conversion among Y-chromosomal duplicate genes or selective constraint (*30-32*). Among the re-sequenced 382 chromosomes with *b2/b3* deletions, no pathogenic mutations were detected in the single retained *BPY2* and *CDY1* gene copies. At the same time, the high rates of large structural rearrangements and copy number variation in the Y chromosome are well established, contrasting with low levels of sequence variation (*19, 33*). One in five or six Estonian Y chromosomes with *gr/gr* and *b2/b3* deletions had undergone secondary rearrangements with no apparent effect on tested andrological parameters and fertility potential (**Table 2**). In the literature, the data about the effects of secondary duplications after an initial *AZFc* partial deletion on sperm parameters are inconclusive. Some studies have suggested increased pathogenicity (*34-38*), whereas others have reported neutral or even positive effects on spermatogenesis (*17, 39-41*). However, further copy number reductions in this genomic region appear to be very rare - none of the 44 *gr/gr* or 631 *b2/b3* deletion carriers were identified with further reductions beyond what is expected from the initial deletion.

In summary, we have undertaken a comprehensive study of the carriers of *AZFc* partial *gr/gr* and *b2/b3* deletions, and uncovered high levels of structural variation in the *AZFc* locus, but low sequence diversity of the coding genes within the region. As a major finding, we discovered a large inversion specific to the Y lineage R1a1-M458 that represents a hotspot for subsequent *AZFc* partial deletions. Men carrying Y chromosomes with this complex rearrangement have >10-fold increased risk of severe spermatogenic failure, but the consequences of this risk could be potentially alleviated by early identification of the variant carriers and facilitating the storage of their sperm samples. Our study results thus have the potential to improve clinical diagnostics and management of idiopathic impaired spermatogenesis in a significant fraction of men originating from Northern and Central European populations.

## Subjects and Methods

### Ethics statement

The study was approved by the Ethics Review Committee on Human Research of the University of Tartu, Estonia (permissions 146/18, 152/4, 221/T-6, 221/M-5, 272/M-13, 267M-13, 286M-18, 288M-13), and sequencing/genotyping was approved at the Wellcome Sanger Institute under WTSI HMDMC 17/105. Written informed consent for evaluation and use of their clinical data for scientific purposes was obtained from each person prior to recruitment. All procedures and methods have been carried out in compliance with the guidelines of the Declaration of Helsinki.

### Study subjects

Patients with idiopathic spermatogenic impairment (n=1,190) were recruited at the Andrology Centre at Tartu University Hospital (AC-TUH) in 2003-2015 (PI: M. Punab). Included cases showed reduced sperm counts (<39 x 10^6^/ejaculate) in at least two consecutive semen analyses (*27*). Recruitment and sampling, semen analyses and hormone assays and definition of idiopathic cases have previously been described in detail (*3*). Men with known causes of male infertility detected during routine diagnostic workup were excluded, e.g. cryptorchidism, testicular cancer, orchitis/epididymitis, mumps orchitis, testis trauma, karyotype abnormalities and complete Y-chromosomal microdeletions. The final idiopathic infertility group included 104 azoospermia (no sperm), 88 cryptozoospermia (sperm counts >0–1 x 10^6^/ejaculate), 319 severe oligozoospermia (1–10 x 10^6^) and 679 moderate oligozoospermia (10–38 x 10^6^) cases (**Table 1, Table S1**).

The reference sample of Estonian men (n=1,134) comprised healthy young men (n=499) and subjects with proven fatherhood (n=635) (**Table 1, Table S1**). The cohort of ‘Estonian young men’ (n=499) was recruited at the AC-TUH in 2003-2004 (PI: M. Punab), representing a healthy male group with median age 18.6 (17.2-22.9) years at the time of recruitment (*42*). The subgroup of ‘Partners of pregnant women’ (n=324) includes male partners of pregnant women, recruited in 2010-2014 at the Tartu University Hospital and the West Tallinn Central Hospital (*3*). All participants in these subgroups were offered complete andrological workup at the AC-TUH; thus, 810 men (of 823) underwent sperm analysis. Further clinical phenotyping details are provided in **Supplemental Data**.

The subgroup of ‘REPROMETA proven fathers’ (n=311) was recruited in 2006-2011 at the Women’s Clinic at Tartu University Hospital during the REPROMETA study (PI: M. Laan), originally designed to collect mother-father-placenta trios at delivery to investigate genetics of pregnancy complications (*43, 44*). In this study, the REPROMETA fathers represented reference men with proven fertility. Only self-reported age and BMI data were available for this subgroup.

### Genotyping Y-chromosomal microdeletions

All study subjects (n=2,324) were typed for complete *AZFa* (loss of markers sY84 and sY86), *AZFb* (sY127 and sY134), *AZFc* (sY254 and sY255), and partial *AZFc* deletions *gr/gr* (sY1291), *b2/b3* (sY1191) and *b1/b3* (sY1161, sY1191 and sY1291) following established multiplex PCR protocols and PCR primers (*7, 45*) (**Table S15**).

### Re-sequencing of retained *DAZ, BPY2* and *CDY* genes using Illumina MiSeq

Re-sequencing of the exonic regions of the retained *AZFc* genes (according to Ensembl release 84) in 476 cases with either *gr/gr* or *b2/b3* deletions targeted in total 94,188 bp per subject. *CDY, BPY2* and *DAZ* genes were amplified using eight, ten or 26 PCR primer pairs, respectively (**Table S16-S17**). The presence of all amplicons was confirmed using gel electrophoresis. Amplicons were pooled in equimolar concentrations, barcoded per sample, and sequenced (250 bp reads, paired-end) on Illumina MiSeq with at least 40 × coverage. BWA (v0.7.15) (*46*) was implemented to map the sequencing reads to a modified human genome reference (GRChg38), where *CDY1a, CDY2a, BPY2a* and either *DAZ3-DAZ4* (*gr/gr* carriers) or *DAZ1-DAZ2* (*b2/b3* carriers) remained unchanged, but the sequences of other *CDY, BPY2* and *DAZ* gene copies were replaced with ‘Ns’. SNVs and indels were identified using GATK HaplotypeCaller (v3.7) with a minimum base quality 20 and outputting all sites (*47*). Y-chromosomal phylogenetic markers were called using bcftools (v1.8) with minimum base quality 20, mapping quality 20 and defining ploidy as 1.

Re-sequencing included 31 patients and 9 reference men with *AZFc gr/gr* deletions. Six men carrying *gr/gr* deletions were not analyzed due to DNA limitations (2 cases) or unavailable andrological data (4 cases). The analysis of *b2/b3* deletion carriers included 382 patients (haplogroup N3: n=380; non-N3, n=2) and 54 ‘Partners of pregnant women’ (N3: n=53; non-N3, n=1).

### Y-chromosomal haplogroup typing

Y lineages of the *gr/gr* samples were defined using 14 markers included in the re-sequencing, plus 34 additional markers determined by Sanger sequencing or restriction fragment length polymorphism (RFLP) analysis (**Table S17-S18**). The *b2/b3*-deletion carriers were typed for Y marker N3-M46 (Tat) (*48*). The sub-lineages of the re-sequenced haplogroup N3 samples were defined in more detail using 16 phylogenetic markers from the Illumina MiSeq dataset, following established nomenclature (*22, 49*). For the other haplogroups, nomenclature according to the International Society of Genetic Genealogy (ISOGG, version 14.14) was followed.

### Determination of *DAZ, BPY2* and *CDY* gene dosage and gene types

The Bio-Rad QX 200 Droplet Digital PCR system was used to quantify the copy numbers of the retained *DAZ, BPY2* and *CDY* genes for 44 *gr/gr* deletion carriers (31 cases, 13 reference men with sperm analysis data) and 631 *b2/b3* carriers (382 cases/249 reference men). PCR primers and probes for gene copy dosage detection are provided in **Table S19** and methodological details in **Supplemental Data**. The re-sequencing data of the *DAZ* genes covered nine paralogous sequence variants (PSVs) that were used to determine the retained gene copies in the *gr/gr* and *b2/b3* deletion carriers (**Table S20**). For the validation of the *DAZ* gene copy mapping approach, at least five *gr/gr* carriers were additionally typed for published SNV combinations differentiating the *DAZ* gene copies (*23, 50*). The retained *CDY1* gene was identified according to Machev et al. (*23*).

### Genetic association testing with andrological parameters

Statistical testing for the associations between *AZFc gr/gr* or *b2/b3* deletions and andrological parameters was conducted using RStudio (version 1.2.1335) and data were visualised using ggplot2 (version 3.2.1) (*51*). Differences in continuous clinical variables between groups were compared using the non-parametric Pairwise Wilcoxon Rank Sum Test. Genetic association with the carrier status of *b2/b3* deletion and its subtypes was also tested using linear regression analyses (**Supplemental Data**).

## Data Availability

Illumina MiSeq re-sequencing data are available through the European Genome-phenome Archive (EGA, https://www.ebi.ac.uk/) under the accession number: EGAS00001002157

https://www.ebi.ac.uk/ega/home

## Supplemental Data

Supplemental Data include Supplemental Subjects and Methods, four figures and 20 tables.

## Acknowledgements

We thank all the patients for making this study possible. The clinical team at the Andrology Centre, Tartu University Hospital is thanked for the professional phenotyping and assistance in patient recruitment over many years. Mart Adler and Eve Laasik are specifically acknowledged for the management of the Androgenetics Biobank, and all M.L. team members are thanked for their contributions to the DNA extractions. Our work was supported by the Wellcome Trust (098051). P.H. and L.K. were supported by Estonian Research Council Grant PUT1036 and M.P. and O.P. by PUT181. L.K., M.G., K.R. and M.L. were supported by Estonian Research Council Project IUT34-12, R.F. by IUT24-1 and S.R. by IUT24-1 and MOBTT53. Establishment of the cohort of Estonian infertility subjects was also supported by the EU through the European Regional Development Fund, project HAPPY PREGNANCY, no. 3.2.0701.12-004 (M.L., M.P., K.R.).

## Author contributions

P.H. conceived and M.P., Y.X., C.T-S. and M.L. supervised the study. M.P. lead, O.P. and P.K. contributed critically to the patient recruitment, phenotyping and clinical data documentation. P.H., L.K. and E.A. performed experiments, P.H., L.K. and M.L. performed analyses. M.G., S.R. and R.F. provided data and assisted with analyses. M.P., K.R., K.M. and M.L. provided DNA samples. P.H., C.T-S., Y.X., M.L. and M.A.J. provided resources. P.H. and M.L. drafted the manuscript with contributions from all other authors. All authors contributed to the final interpretation of data and critical reading of the manuscript.

## Web resources

International Society of Genetic Genealogy (ISOGG) https://isogg.org

RStudio: http://www.rstudio.com/

Primer3plus: https://primer3plus.com

Ensembl: https://www.ensembl.org/

Variant Effect Predictor tool (VEP): https://www.ensembl.org/Tools/VEP

## Supplemental Subjects and Methods

### Details on the clinical assessment of the study subjects

All men, who had turned to Andrology Centre, Tartu University Hospital (AC-TUH) due to idiopathic infertility (n=1,190), as well as the participants of the ‘Estonian young men’ cohort (n=499) and the subgroup ‘Partners of pregnant women’ (n=324) were offered complete routine andrological workup. The subjects were examined by specialist andrologists at the AC-TUH, who had received respective training in clinical assessment and standardized andrological workup, locally and in collaboration with other European Andrology Academy (EAA) accredited centers. Also, anthropometric parameters were documented during clinical examination. Details are described in (*1*).

Physical examination for the assessment of genital pathology and testicular size (orchidometer; made of birch wood, Pharmacia & Upjohn, Denmark) was performed with the patients in standing position. The total testes volume is the sum of right and left testicles. The position of the testicles in the scrotum, pathologies of the genital ducts (epididymis and ductus deference) and the penis, urethra, presence and if applicable grade of varicocele were registered for each subject.

For 2,000 study subjects sperm analysis was performed, whereas 13 reference cases did not agree with this procedure. Semen samples were obtained by patient masturbation and semen analysis was performed in accordance with the World Health Organization (WHO) recommendations. In brief, after ejaculation, the semen was incubated at 37°C for 30–40 min for liquefaction. Semen volume was estimated by weighing the collection tube with the semen sample and subsequently subtracting the predetermined weight of the empty tube assuming 1 g = 1 mL. For assessment of the spermatozoa concentration, the samples were diluted in a solution of 0.6 mol,L NaHCO3 and 0.4% (v,v) formaldehyde in distilled water. The spermatozoa concentration was assessed using the improved Neubauer haemocytometers.

Genomic DNA was extracted from EDTA-blood. After blood draw in the morning, serum and plasma fractions were separated immediately for hormone measurements (FSH, LH, testosterone). All laboratory analyses and routine genetic testing (karyotyping, Y-chromosomal microdeletions) were performed at the United Laboratories of Tartu University Hospital according to the established clinical laboratory guidelines. Detailed methodology and reference values for hormonal levels are available by the service provider: https://www.kliinikum.ee/yhendlabor/analueueside-taehestikuline-register.

### Analysis of variant effects from the Illumina MiSeq dataset

Variant effect prediction was performed using the Variant Effect Predictor tool (VEP, https://www.ensembl.org/Tools/VEP, Ensembl release 99) (*2*). The Combined Annotation Dependent Depletion (CADD) score ≥ 20, i.e., including variants among the top 1% of deleterious variants in the human genome, was considered indicative of potential functional importance of identified SNVs in the coding regions (*3*).

### *DAZ, BPY2* and *CDY* copy number detection using Droplet Digital PCR

The PCR primers and probes were designed using Primer3plus (version 2.4.2), PCR reactions were performed according to the recommendations in the Droplet Digital PCR Application Guide (Bio-Rad, U.S.) (**Table S19**) and described in (*4*). A XQ200 Droplet Reader was used to measure the fluorescence of each droplet and QuantaSoft software (v1.6.6.0320; Bio-Rad) to cluster droplets into distinct fluorescent groups. The copy number of each gene was determined by calculating the ratio of target (unknown - *DAZ, BPY2* or *CDY*) and reference (single-copy *SRY* gene) concentration.

ddPCR reactions for each gene were performed once for every sample. For samples carrying the *gr/gr* deletion, if the copy number obtained differed from the expected (two copies of *DAZ* and *BPY2*, three copies of *CDY*), then the ddPCR reaction was repeated. For *b2/b3* carriers typing was repeated for all samples not carrying the two most typical copy numbers (2-1-3 or 4-2-4 copies of *DAZ, BPY2* and *CDY* genes, respectively). Additionally, a total of 5% of random samples were replicated. If the copy number estimates between replicates differed by 0.8 or more, then a third replicate was performed, and the final copy number was calculated as average of the two closest replicates.

### Genetic association analysis using linear regression

Genetic association with the carrier status of *b2/b3* deletion and its subtypes was also tested using linear regression analyses adjusted for age. For sperm parameters abstinence time and for total testosterone levels BMI estimates were additionally used as cofactors. Natural log transformation was used to achieve an approximately normal distribution of values. In all cases (except total sperm counts), the applied transformation resulted in a close-to-normal distribution of values. For the linear regression analyses, statistical significance threshold after correction for multiple testing was estimated *P*<1.0 x 10^−3^ (6 tests x 8 independent parameters).

### Protocol for the detection of *AZFc* rearrangement at the Y lineage R1a1a1b1a1a-M458: *r2/r3* inversion followed by partial deletions

#### Step 1. Multiplex PCR for typing the *AZFc* partial deletions (modified from (*5, 6*))

The most commonly used method to detect *AZFc* partial deletions is the plus/minus STSs-based PCR assay that detects the presence of five markers around *AZFc* region (Figure 1B, Table S2). The multiplex PCR reactions contained the final concentrations of 1X PCR buffer B1 (Solis Biodyne, Estonia), 2.5 mM MgCl_2_, 2.5 mM dNTP, 2 µM PCR primers for STS markers sY1291 and sY1201, 3 µM PCR primers for STS markers sY1191, sY1206 and sY1161 (**Table S15**), 1U FIREPol^®^ DNA polymerase (Solis Biodyne) and 10 ng of template genomic DNA per reaction. The following PCR conditions were used: for 5 min at 95°C, followed by 32 cycles of 30 sec at 95°C, 30 sec at 63°C and 1 min at 72°C, final extension of 10 min at 72°C and a 4°C hold. The presence/absence of PCR products in a reaction were checked on 2% agarose gel. Lack of amplification of STS marker sY1291 (but presence of all others) was used to determine the *gr/gr* deletion, and lack of sY1191 the *b2/b3* deletion.

#### Step 2. Analysis of the allelic state of Y lineage R1a1a1b1a1a-specific phylogenetic marker M458

The phylogenetic marker M458 (rs375323198, A > G polymorphism, GRCh38 genomic coordinate: chrY: 22220317) indicating the carrier status of *r2/r3* inverted Y chromosome was amplified using the following conditions: PCR reactions contained the final concentrations of 1X PCR buffer B1 (Solis Biodyne), 2.5 mM MgCl_2_, 2.5 mM dNTP, 10 µM forward and reverse PCR primers for M458 (see **Table S18** for primer sequences), 1U FIREPol^®^ DNA polymerase (Solis Biodyne OÜ) and 10 ng of template genomic DNA per reaction. The following PCR conditions were used: for 5 min at 95°C, followed by 32 cycles of 30 sec at 95°C, 30 sec at 52°C and 1 min at 72°C, final extension of 10 min at 72°C and a 4°C hold. The presence of the M458 marker in derived state (instead of the ancestral allele ‘A’ presence of allele ‘G’ at position 147) was determined using Sanger sequencing.

#### Step 3. Analysis of retained *DAZ* gene copies by typing gene-specific PSVs (*optional*)

The retained *DAZ* gene copies can be identified by typing gene copy-specific paralogous sequence variants (PSVs). In the current study, the retained *DAZ* copies were identified by determining the allelic states of 9 PSVs from the Illumina MiSeq re-sequencing data (see **Table S20** for full details). Additionally, a number of published protocols are available, for full details see (*7-9*).

**Figure S1.**
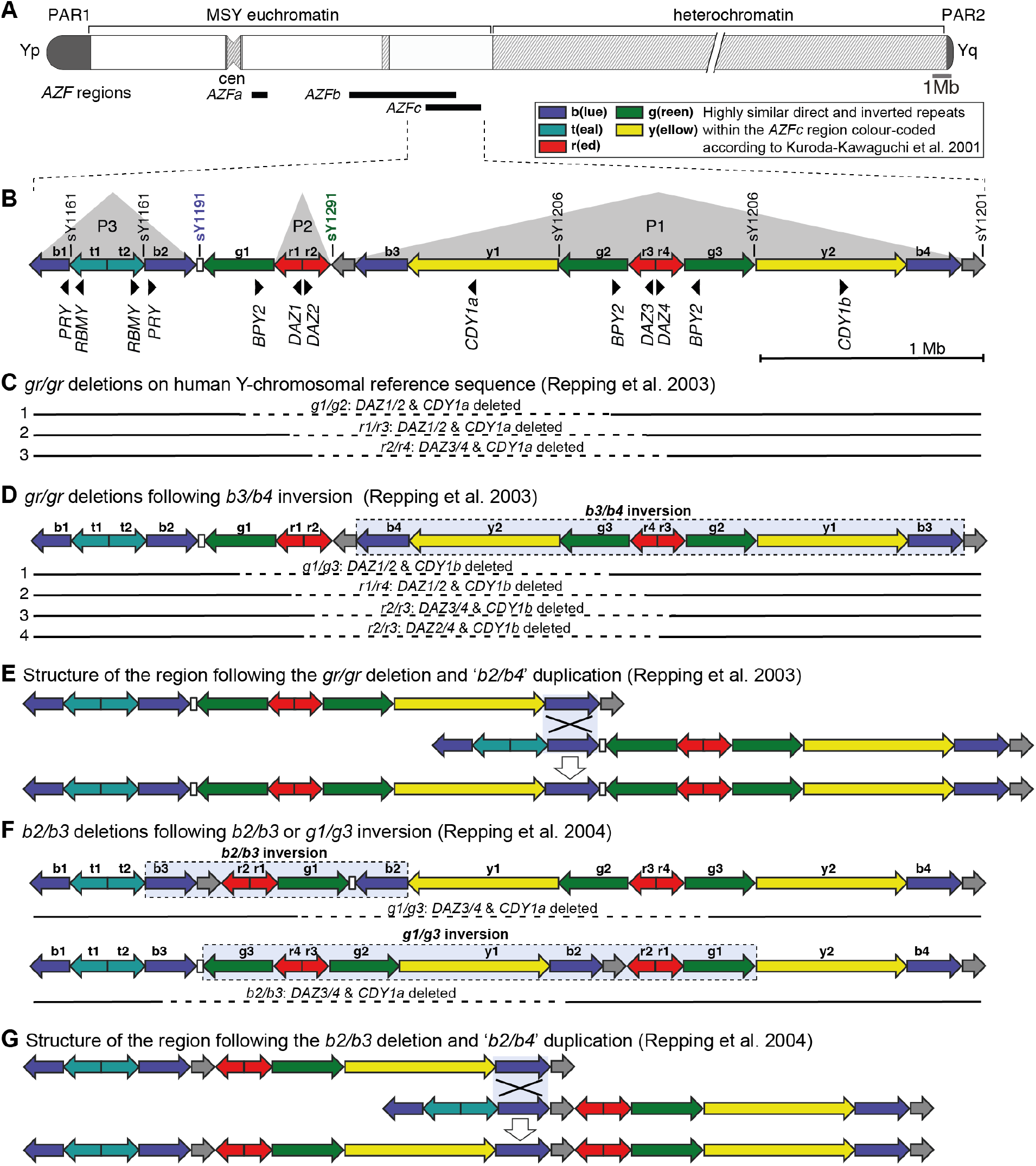
The human Y chromosome and the *Azoospermia Factor (AZF*) regions. **(A.)** The human Y chromosome drawn to approximate scale with regions of *AZFa, AZFb* and *AZFc* deletions shown below as black bars. PAR - pseudoautosomal region, MSY - male-specific region of the Y chromosome, cen - centromere, *AZF* - azoospermia factor region. **(B.)** Structure of the *AZFc* region on human Y-chromosomal reference sequence with approximate locations of protein-coding genes and STS (sY) markers used for detection of partial *AZFc* deletions (*10*). Alternative involved regions in *gr/gr* deletions arising on (**C.)** the human Y-chromosomal reference sequence and (**D.)** the Y chromosome with a preceding *b3/b4* inversion. The approximate region removed by each deletion is shown as a dashed line. **(E.)** Structure of the *AZFc* region undergone the *gr/gr* deletion and the proposed model of homologous recombination leading to the subsequent ‘*b2/b4*’ duplication. The light blue box denotes the recombination targets. The duplication is presumably the result of recombination between sister chromatids. **(F.)** Reported models for *b2/b3* deletions arising on the Y chromosomes with preceding *b2/b3* or *g1/g3* inversions. The approximate region removed by each deletion is shown as a dashed line. **(G.)** Structure of the *AZFc* region undergone the *b2/b3* deletion and the proposed model of homologous recombination leading to the subsequent ‘*b2/b4*’ duplication. The light blue box denotes the recombination targets. The duplication is presumably the result of recombination between sister chromatids.

**Figure S2.**
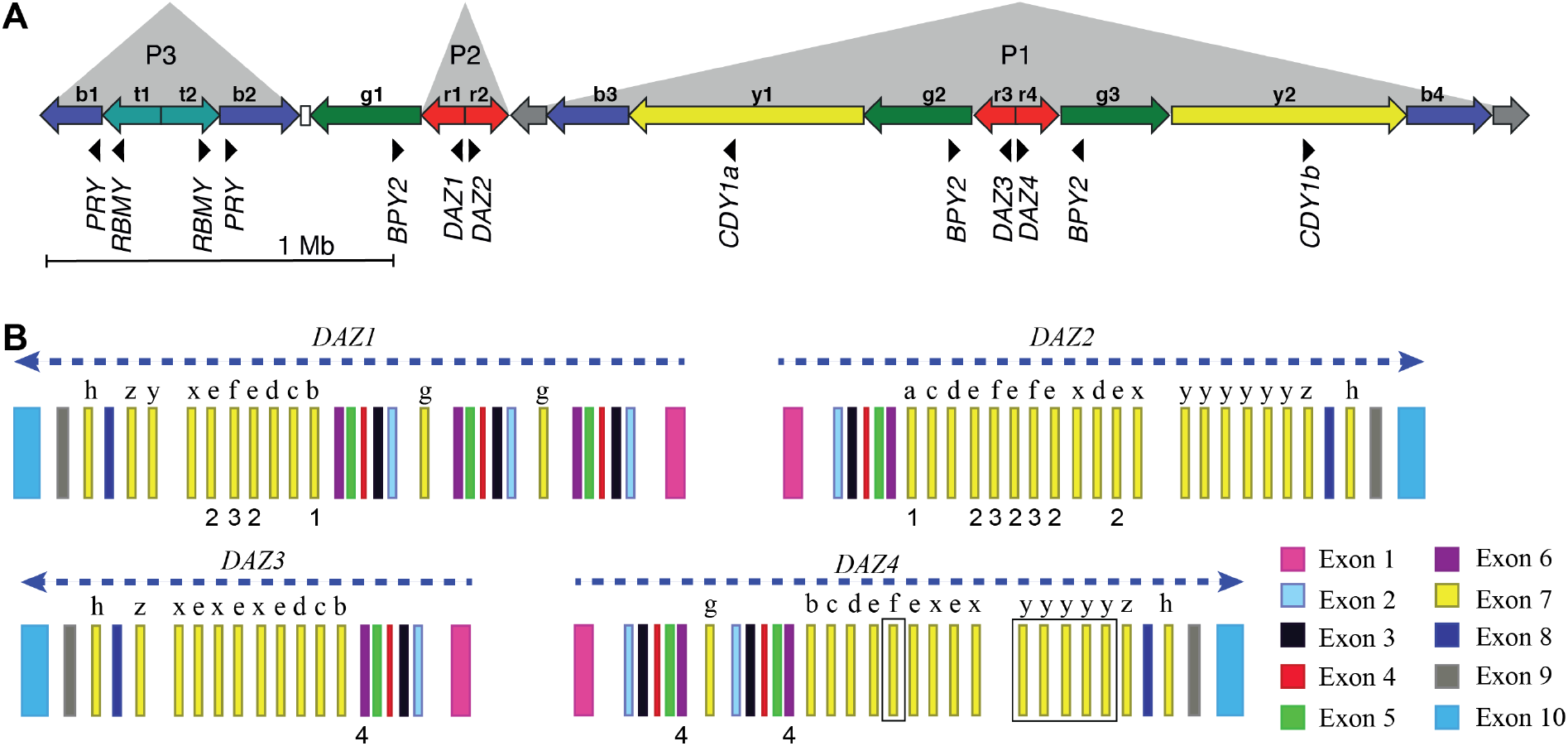
Location of the identified exonic variants in the *DAZ* genes. **(A.)** Schematic representation of the human Y-chromosomal reference sequence containing the *AZFc* region, with approximate locations of protein-coding genes and their direction of transcription shown as black triangles. Direct and inverted repeats with highly similar DNA sequences are denoted as coloured arrows (b - blue, t - teal, g - green, r - red, y - yellow as originally described by (*10*); arrow direction indicates the orientation of the gene from transcription start site. **(B.)** The structure of human *DAZ* genes, modified from (*7*) to represent the human Y-chromosomal reference sequence (GRCh38). Blue dashed arrows show the direction of transcription. Exons with high DNA sequence similarity within and between *DAZ* genes are denoted with the same fill colour. The number of highly similar exon 7 copies (in yellow) varies between genes and copies marked with the same letter denote identical sequences. Numbers below the gene structure denote exonic variants: 1 - DAZ1 p.H173Y or DAZ2 p.H173Y 2 - DAZ1 p.Q262E or DAZ2 p.Q262E 3 - DAZ1 p.Y243C or DAZ2 p.Y219C 4 - potential splicing variant in *DAZ3* or *DAZ4*. Black squares in *DAZ4* denote the deleted exons 7f and 7y in Y lineages I1 and R1a1a1b1a2, respectively.

**Figure S3.**
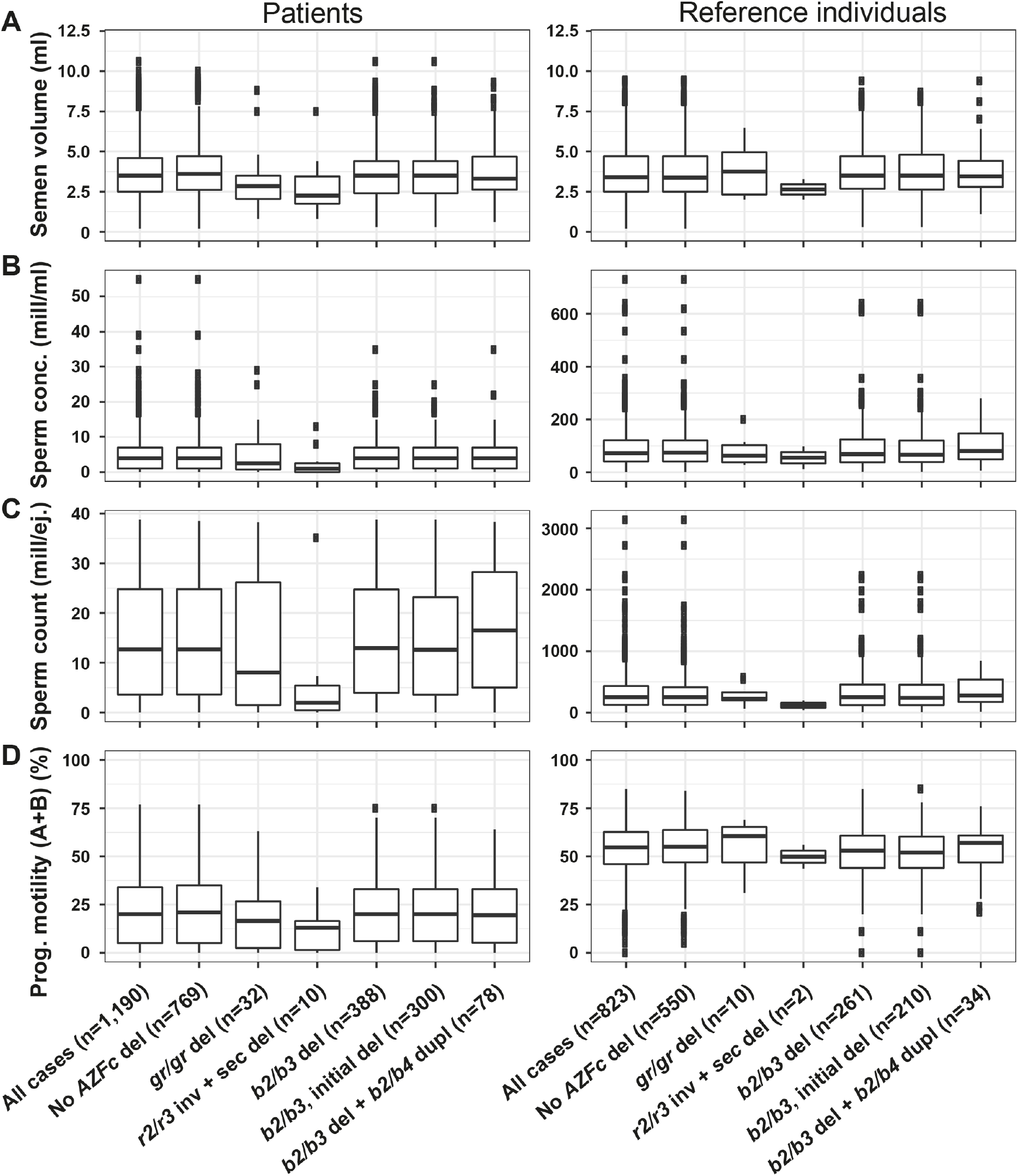
Distribution of seminal parameters in idiopathic male factor infertility cases with spermatogenic impairment and reference subjects. Samples are grouped by genotype with the number of samples (n) shown in brackets. del - deletion, sec - secondary, inv - inversion, dupl - duplication, mill - million, ej. - ejaculate. Note different scaling of the Y-axis for the two study groups in panels B and C.

**Figure S4.**
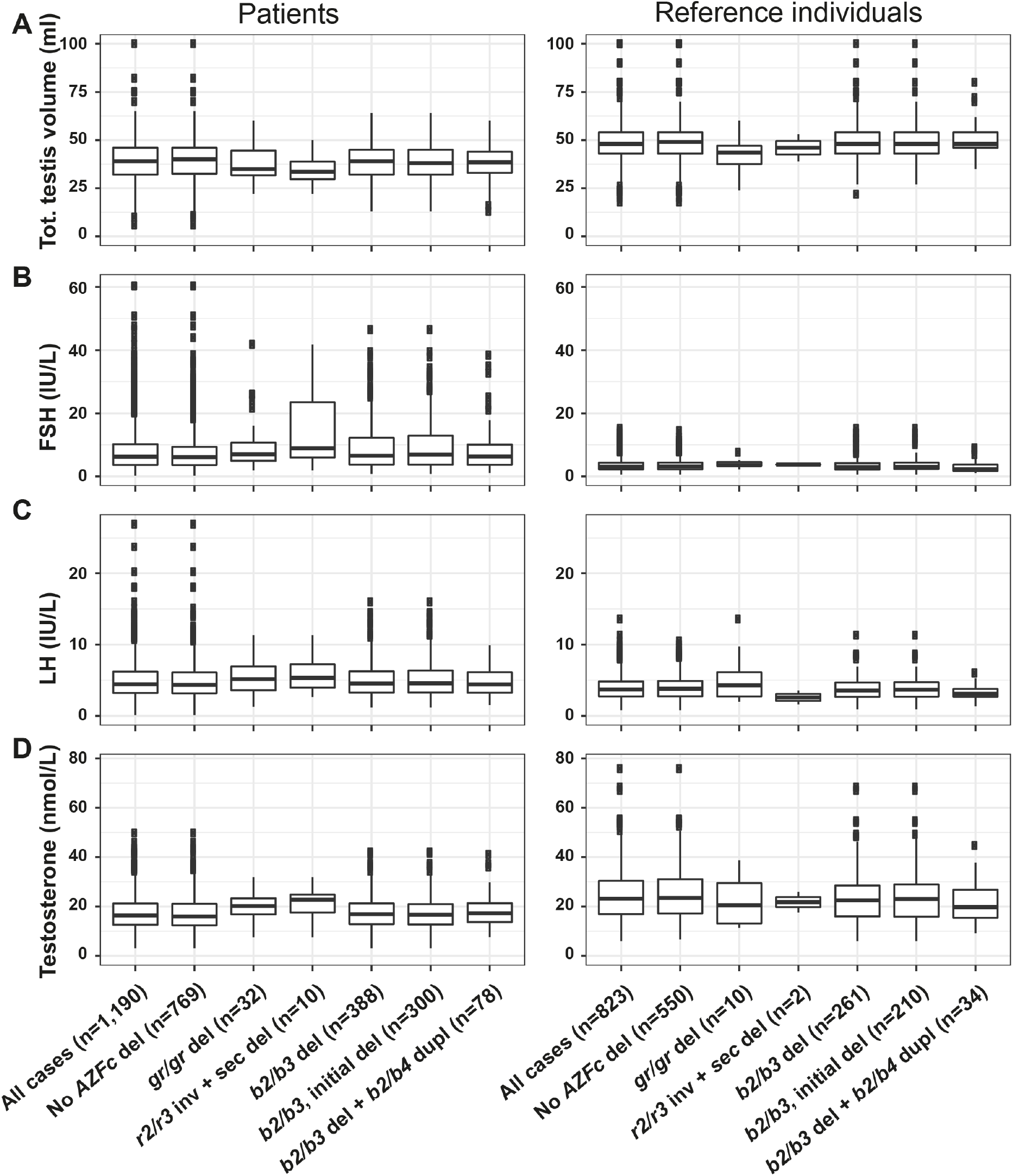
Distribution of hormonal and testicular parameters in idiopathic male factor infertility cases with spermatogenic impairment and reference individuals. Samples are grouped by genotype with the number of samples (n) shown in brackets. del - deletion, sec - secondary, inv - inversion, dupl - duplication.

